# Evidence of co-infection during Delta and Omicron variants of concern co-circulation, weeks 49-2021 to 02-2022, France

**DOI:** 10.1101/2022.03.02.22271694

**Authors:** Patricia Combes, Maxime Bisseux, Antonin Bal, Pierre Marin, Christine Archimbaud, Amélie Brebion, Hélène Chabrolles, Christel Regagnon, Jérémy Lafolie, Gregory Destras, Bruno Simon, Laurence Josset, Cécile Henquell, Audrey Mirand

**Affiliations:** CHU Clermont-Ferrand, pôle BMAP, F-63003 Clermont-Ferrand, France; CHU Clermont-Ferrand, 3IHP, Virology laboratory, National Reference Centre for Enteroviruses and parechoviruses-associated laboratory, F-63003 Clermont-Ferrand, France; Université Clermont Auvergne, CNRS 6023-LMGE, EPIE, F-63001 Clermont-Ferrand, France; GenEPII Sequencing Platform, Institut des Agents Infectieux, Hospices Civils de Lyon, F-69004 Lyon, France; CIRI, Centre International de Recherche en Infectiologie, Team VirPath, Univ Lyon, Inserm U1111, Université Claude Bernard Lyon 1, CNRS, UMR5308, ENS de Lyon, F-69007 Lyon, France; CH Vichy, Laboratory Department, F-03000 Vichy, France

**Author notes:** **Corresponding author:** Audrey Mirand.

**Keywords:** SARS-CoV-2 co-infection, Delta and Omicron VOC, Single nucleotide polymorphism screening, Whole genome sequencing

## Abstract

We report evidence of Delta/Omicron SARS-CoV-2 co-infections during the fifth wave of COVID-19 pandemics in France for 7 immunocompetent and epidemiologically unrelated patients. These co-infections were detected by PCR assays targeting SARS-CoV-2 S-gene mutations K417N and L452R and confirmed by whole genome sequencing which allowed the proportion estimation of each subpopulation. For 2 patients, the analyses of longitudinal samples collected 7 to 11 days apart showed that Delta or Omicron can outcompete the other variant during dual infection.

---

The 21K/L Omicron variant of SARS-COV-2, classified as a Variant of Concern (VOC) was first described in November 2021 in South Africa. In Europe, as elsewhere, Omicron co-circulated with the 21J Delta VOC which it rapidly displaced [1]. During this period of intense viral co-circulation, we identified co-infection of the Omicron and Delta VOC in 7 immunocompetent patients.

## Co-infection Delta/Omicron detection

After RNA extraction with the MGI Nucleic Acid Extraction Kit on the automated MGI-SP960 platform (BGI, Shenzhen, China), SARS-CoV-2 genome was detected in respiratory samples by RT-PCR with the TaqPath™ COVID-19 RT-PCR Kit (ThermoFisher Scientific, Massachusetts, USA) which targets three genes (ORF1ab, N and S) on a QuantStudio5 thermocycler, according to the manufacturer’s instructions. Based on S-gene target failure (SGTF), this kit allows pre-screening of variants such as 20I Alpha and 21K Omicron, which carry the Δ69-70 deletion [2,3]. In accordance with national French recommendations in December 2021 [4], SARS-CoV-2 positive samples were further screened for VOC-specific amino-acid substitution by targeting the single nucleotide polymorphisms (SNPs) S:L452R and S:K417N with the TaqMan™ SARS-CoV-2 mutation panel molecular assay (ThermoFisher Scientific) to rapidly detect 21I/J Delta and 21K/L Omicron, respectively. Amplification curves and genotyping results were analysed with the QuantStudio Design and Analysis 2.4 software which discriminates between wild type and mutated samples. The 21K Omicron was first detected at the university hospital of Clermont-Ferrand, France on 6 December 2021 (week 49). It co-circulated with Delta and their respective proportions have varied inversely since (Figure 1). Between weeks 49-2021 and 02-2022, SARS-CoV-2 genome was detected in 3,831 respiratory samples, of which 3,237 (84.5%) were screened for VOC specific SNPs. Unexpected mutation profiles were observed in 7 nasopharyngeal samples (0.2%) as both S:K417N and S:L452R mutations were detected suggesting a dual Delta/Omicron population (Figure 2). None of these samples showed the characteristic S gene dropout signature of the variant 21K Omicron with the RT-PCR kit.

**Figure 1.**
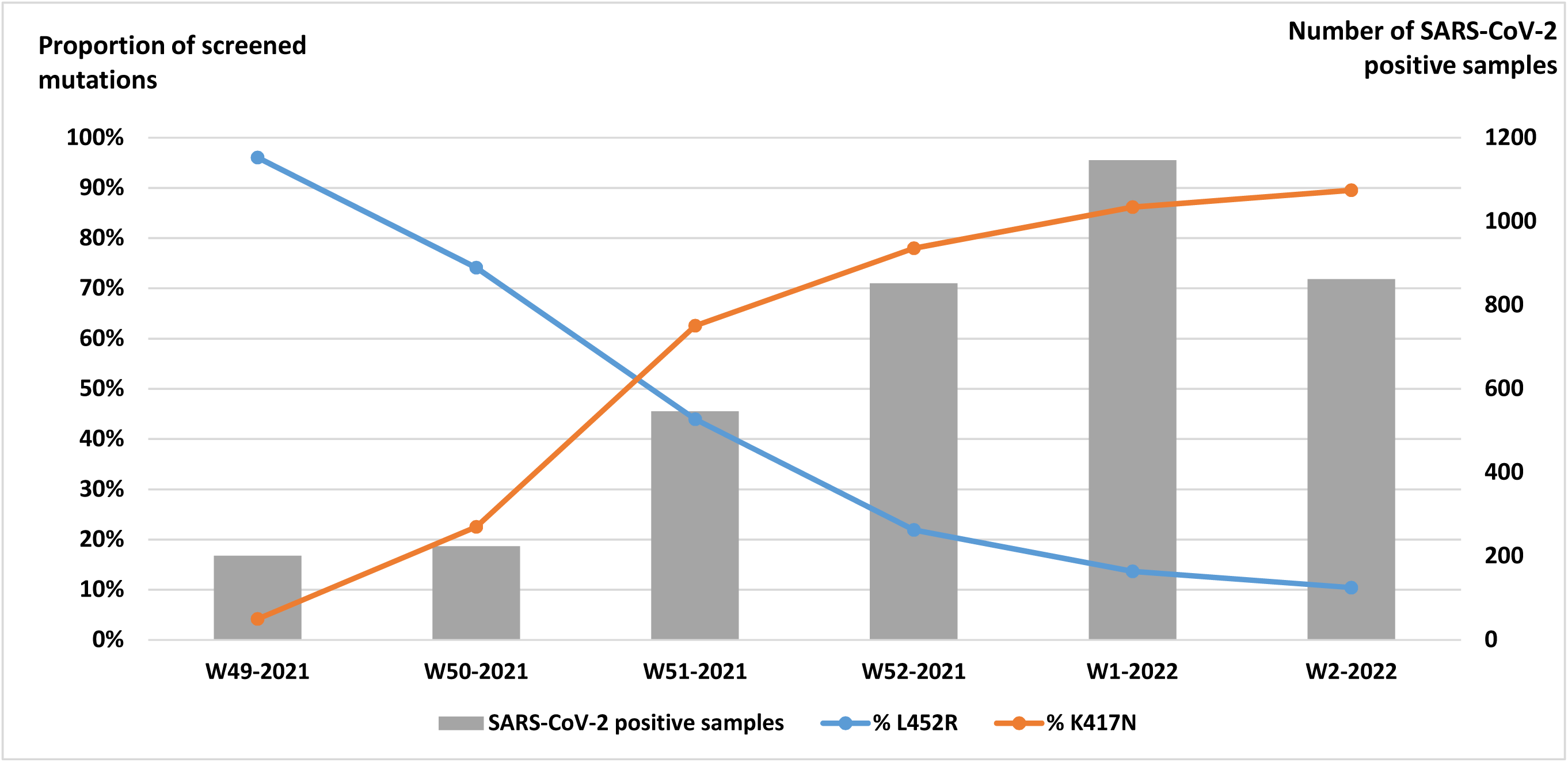
Weekly distribution of S:L452R and S:K417N single nucleotide polymorphism detection on 3,237 SARS-CoV-2 positive screened samples from weeks 49-2021 to 02-2022, University Hospital of Clermont-Ferrand, France.

**Figure 2.**
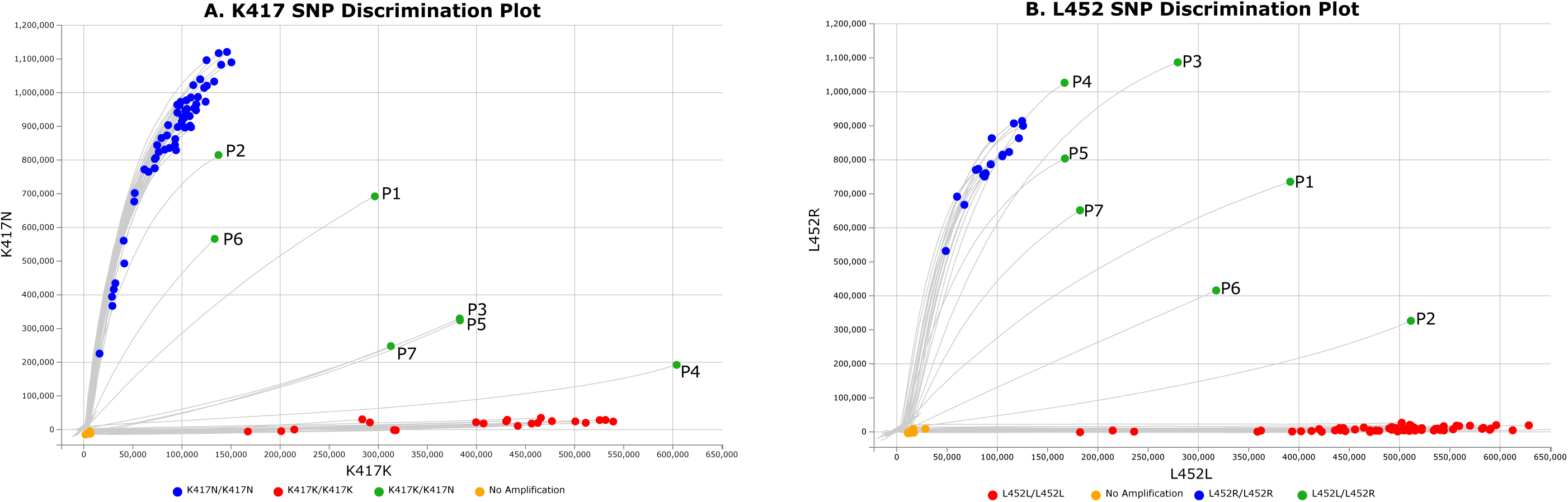
Screening profiles for VOC-specific amino-acid substitution S:L452R and S:K417N suggestive of a dual population Screening was performed with the TaqMan™ SARS-CoV-2 mutation panel molecular assay (ThermoFisher Scientific, Massachusetts, USA). For each mutation of interest, probes labelled with VIC or FAM detect the wild type (WT) and the mutated sequence, respectively. WT samples are represented as red dots along the x-axis and mutated samples as blue dots along the y-axis. A dual population is suspected by intermediate mutation profile with detection of WT and mutated sequences in the same sample and is represented by green dots. Abbreviations: SNP, Single Nucleotide Polymorphism

### Clinical characteristics of patients with suspected co-infection Delta/Omicron

The seven patients were non immune-compromised adults. Six were outpatients under 35 years old presenting with mild infection (n=5) or asymptomatic infection (n=1) and all but one had received 2 or 3 doses of vaccine. The last patient (P6) was over 70 years old, unvaccinated and required hospitalisation for hypoxemic respiratory failure 9 days after the diagnosis of infection (Table 1). Follow-up samples collected 11 (D11) and 7 days (D7) after the initial diagnosis (D1 samples) were obtained for patients P1 and P5, respectively.

**Table 1.** Demographics, clinical and virological characteristics of patients with co-infection by Delta and Omicron.

| # Patient | P1 |  | P2 | P3 | P4 | P5 |  | P6 | P7 |
| --- | --- | --- | --- | --- | --- | --- | --- | --- | --- |
| Age group (years) | 31-40 |  | 21-30 | 31-40 | 11-20 | 21-30 |  | 71-80 | 21-30 |
| Sex | M |  | M | M | M | F |  | F | M |
| COVID-19 vaccination | yes |  | yes | yes | no | yes |  | no | yes |
|  | 2 doses of BNT162b2, last one in April 2021 |  | 2 doses of BNT162b2, last one in July 2021 | 3 doses of mRNA-1273, last dose one week before the diagnosis |  | 2 doses of BNT162b2 + 1 dose of mRNA-1273 in December 2021 |  |  | 2 doses of BNT162b2, last one in June 2021 |
| Symptoms | Mild |  | Mild | Mild | Mild | Asymptomatic |  | Severe | Mild |
| Sampling week <sup>e</sup> | D1<br>W50-2021 | D11<br>W52-2021 | D1<br>W51-2021 | D1<br>W51-2021 | D1<br>W1-2022 | D1<br>W1-2022 | D7<br>W2-2022 | D1<sup>f</sup><br>W2-2022 | D1<br>W51-2021 |
| RT-PCR (Ct) <sup>a</sup> |  |  |  |  |  |  |  |  |  |
| ORF1ab | 18.18 | 30.59 | 17.39 | 22.25 | 17.06 | 27.38 | 31.84 | 27.53 | 21.77 |
| N | 18.04 | 30.58 | 17.22 | 21.15 | 14.74 | 28.11 | 31.85 | 28.54 | 19.38 |
| S | 19.73 | 29.99 | 20.86 | 23.60 | 17.46 | 30.68 | SGTF | 26.18 | 23.43 |
| SNP screening K417N <sup>b</sup> | yes | no | yes | yes | yes | yes | yes | yes | yes |
| SNP screening L452R <sup>b</sup> | yes | yes | yes | yes | yes | yes | no | yes | yes |
| WGS, % of non-N bases > 10x | 99.72% | 97.68% | 99.51% | 98.19%<sup>h</sup> | 99.69% | 99.7% | 98.01% | 99.12% | 78.15%<sup>h</sup> |
| WGS, coverage | 6728x | 3363x | 7625x | 4225x<sup>h</sup> | 4217x | 6582x | 2896x | 5992x | 475x<sup>h</sup> |
| Lineage <sup>c</sup> | BA.1 | AY.122 | BA.1.1 | AY.127<sup>h</sup> | AY.4.10 | AY.122 | BA.1 | None | n.i.<sup>i</sup> |
| Clade <sup>d</sup> | 21K<br>(Omicron) | 21J<br>(Delta) | 21K<br>(Omicron) | 21J<sup>h</sup><br>(Delta) | 21J<br>(Delta) | 21J<br>(Delta) | 21K<br>(Omicron) | 21K<br>(Omicron) | n.i. |
| Private mutations score <sup>d</sup> | 450 | 29 | 108 | 88<sup>h</sup> | 0 | 121 | 67 | 704 | n.i. |
| **Number of clade defining mutations** |  |  |  |  |  |  |  |  |  |
| 36 Delta (D) mutations | 36/36 (D) | 36/36 (D) | 32/36 (D) | 34/36<sup>h</sup> (D) | 36/36 (D) | 36/36 (D) | 5/36 (D) | 34/36 (D) | n.i. |
| 72 Omicron (O) mutations | 72/72 (O) | 29/72 (O) | 72/72 (O) | 59/72<sup>h</sup> (O) | 35/72 (O) | 69/72 (O) | 66/72 (O) | 59/72 (O) |  |
| **P681H/R [% of variant allele frequency]** |  |  |  |  |  |  |  |  |  |
| C23604G, S:P681R Delta (D) | 52% (D) | 100% (D) | 14,2% (D) | 74,6%<sup>h</sup> (D) | 100% (D) | 74,7% (D) | 0% (D) | 11,7% (D) | n.i. |
| C23604A, S: P681H Omicron (O) | 48% (O) | 0% (O) | 85,7% (O) | 25,4%<sup>h</sup> (O) | 0% (O) | 25,3% (O) | 99,8% (O) | 88,2% (O) |  |
| **Clade defining mutations unaffected by primer site mutations, % <sup>g</sup>** |  |  |  |  |  |  |  |  |  |
| Delta (D) | 43.6% ± 11.5% (D) | 96.0% ± 4.8% (D) | 6.8% ± 3.7% (D) | 72.3% ± 12.5%<sup>h</sup> (D) | 94.1% ± 3.7% (D) | 81.8% ± 6.1% (D) | 0% (D) | 57.5% ± 36.6% (D) | n.i. |
| Omicron (O) | 54.2% ± 7.7% (O) | 2.1% ± 3.7% (O) | 90.8% ± 5.7% (O) | 20.2% ± 13.3%<sup>h</sup> (O) | 4.1% ± 3.0% (O) | 17.9% ± 5.2% (O) | 100% (O) | 56.1% ± 32.3% (O) |  |
| SRA PRJNA809680 Accession Numbers | SAMN26197536 | SAMN26197537 | SAMN26197538 | SAMN26197539 | SAMN26197540 | SAMN26197541 | SAMN26197542 | SAMN26197543 | - |
| Conclusion <sup>j</sup> | Co-infection |  | Co-infection | Co-infection | Co-infection | Co-infection |  | Probable recombinant | Probable co-infection |

<sup>a</sup> RT-PCR for SARS-CoV-2 genome detection performed with the TaqPath™ COVID-19 RT-PCR Kit (ThermoFisher Scientific, Massachusetts, USA)
<sup>b</sup> Single nucleotide polymorphism screening was performed with the TaqMan™ SARS-CoV-2 mutation molecular assay K417N and L452R
<sup>c</sup> The lineage was obtained with the Pangolin version v3.1.19 (PangoLEARN release date: 20/01/2022)
<sup>d</sup> The clade and the private mutation scores were obtained with NextClade v1.13.1.
<sup>e</sup> The term D1 (Day 1) is used for the day of diagnosis in our laboratory and the follow up samples days are assessed from the D1 samples.
<sup>f</sup> P6 was diagnosed 9 days earlier the D1 in another laboratory
<sup>g</sup> Artic v4 primer set, unless otherwise indicated
<sup>h</sup> Artic v3 primer set
<sup>i</sup> Interpretable WGS results were for sequences with a median coverage >1000x and a percentage of non-N bases > 95%
<sup>j</sup> The co-infection was classified as probable when it was not confirmed by whole genome sequencing
Abbreviations: Ct, Cycle Threshold; SNP, Single Nucleotide Polymorphism; WGS, Whole Genome Sequencing; n.i., non interpretable; SRA, Sequence Read Archive; SGTF, S Gene target Failure

### Confirmation of co-infections by whole genome sequencing

All samples underwent whole genome sequencing (WGS). Briefly, cDNA was synthesized from two independent RNA extracts and libraries prepared using the COVIDSeq-Test™ (Illumina, San Diego, USA) with the Artic v3 and v4 primer sets following the manufacturer’s guidelines and sequenced with 75 bp paired-end reads on an Illumina NextSeq500 system. Data were processed with the Dragen Lineage pipeline 2.5.4 which provided the Pango lineage, Fasta consensus sequences, bam files and hard-filtered vcf files. Clades were obtained from consensus sequences with Nextclade v1.13.1 (https://clades.nextstrain.org/). Reads were aligned to the reference sequence NC_045512.2. The minimum number of reads to call a base was set at 10 and the minimum Variant Allele Frequency (VAF) to call a mutation for the fasta consensus generation at 0.5. Quality criteria, defined by a median coverage higher than 1000x and a percentage of non-N bases higher than 95% were fulfilled for 8/9 samples (Table 1). Bam and vcf files were analysed to further investigate the presence of dual populations. The presence of co-infection Delta/Omicron was determined by the number of 21J and 21K clade-defining mutations after analysis of the vcf files including major and minor alleles (from the list on https://covariants.org/variants as of 11/02/2022). The proportion estimations of each population were assessed by 1) the VAF at the genomic position 23604 (S:P681) which is unaffected by template-mismatch in the primers and which differs between Omicron (23604A; S:P681H) and Delta (23604G, S:P681R), and 2) the average of the VAF clade-defining mutations. This last parameter was analysed by excluding clade-defining mutations in regions where amplification can be hampered by primer mismatches due to genomic viral mutations with VAF ≥ 5% and not located in the last five nucleotides at the 3’ end of the primers as such mismatches can generate depth dropouts and could promote the amplification of one specific variant [5]. Mutations S:K417N (G22813T) and S:L452R (T22917G) could not be investigated as they are in regions impacted by mutations on primer sites. The raw reads were deposited in the Sequence Read Archive database (reference PRJNA809680). The co-infection was confirmed for 6/7 D1 samples, with both Delta and Omicron variants being detected in varying proportions (Table 1). The two proportion estimation methods produced comparable results except for the sample of P6. The distribution of VAF defining mutations of both variants was homogenous over the whole genome except for the sample of P6, which had predominant VAF Omicron mutations in the 5’part of the genome (Figure 3, Supplementary Figure S1). WGS results for patients P1, P5 and P6 were confirmed with another WGS method using independent RNA extraction and xGen(tm) SARS-CoV-2-Midnight-1200 Amplicon (IDT) sequencing on an Illumina Platform which provided similar results (Supplementary Figure S2).

**Figure 3.**
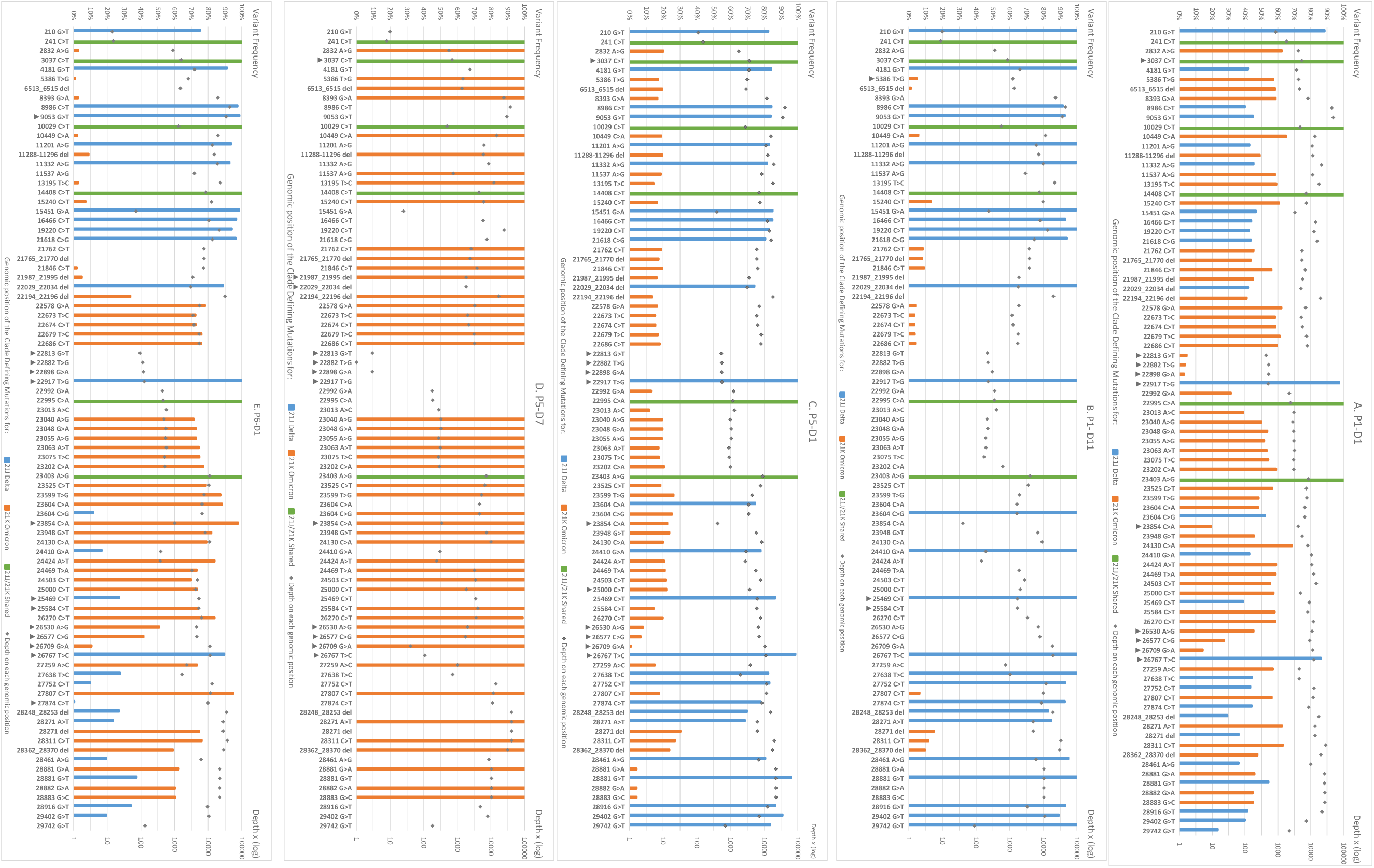
Representation of variant allele frequency clade-defining mutations for 21J Delta and 21K Omicron along the SARS-CoV-2 genome.

All 21J and 21K clade defining mutations along the genome are presented for P1-D1 (A) and P1-D7 (B), P5-D1 (C) and P5-D7 (D) and P6-D1 (E). Mutations are represented with the variant allele frequency on the first Y axis and the depth of each genomic position on the secondary Y axis. If the mutation was not present in the vcf file, the value was set at zero. The black arrows indicate clade defining mutations or deletions excluded for the proportion’s estimation of co-infection (no more than 12 per sample).

### Evolution of the co-infection

Follow-up samples obtained for patients P1 and P5 were analysed by the same methods. For P1-D11 collected 11 days after the first detection, SNP screening showed a mutation profile L452R/K417K suggesting the presence of the Delta variant only. Sequence analysis showed a proportion of Delta-defining mutations at 96.7% with a low proportion of Omicron-defining mutations. For the P5-D7 sample, SNP screening suggested the presence of the Omicron variant (L452L/K471K) and only the Omicron-defining mutations were evidenced by WGS. P1-D1 was co-infected with balanced proportions of Delta and Omicron variants, and our results showed that the Delta variant outcompeted the Omicron variant at D11. In contrast, the Delta variant accounted for 80% of the co-infection in the P5-D1 sample while Omicron variant represented 100 % of the viral population 7 days after. These two follow up samples showed no divergence neither in the number or the percentage of defining mutations all along the genome, suggesting that no recombination occurred between the two strains.

## Discussion

This work shows that co-infection with two SARS-CoV-2 VOC can occur, particularly during a major epidemic wave marked by a co-circulation of two highly transmissible variants in the same region as occurred in December 2021 in France [6]. Few cases of SARS-CoV-2 co-infections have been reported [7–9]. The detection of co-infections could be underestimated by the differences in VOC screening strategies between laboratories and countries. The use of specific RT-PCR assays targeting SNP is an efficient strategy for rapid detection of circulating SARS-CoV-2 VOC and can detect co-infection only if SNPs of different variants are simultaneously screened and if the allele discrimination plots are carefully analysed. In our study, Nextclade assigned a clade from consensus sequences for all sequenced D1 samples but reported the presence of additional mutations compared to the assigned clade as shown by high private mutation scores. Although this score lacks specificity to detect co-infections, it could be a first warning to analyse more carefully the WGS data. The number and prevalence of low-frequency variants for clade defining mutations can be used to detect co-infection but their determination is not systematically analysed in pipelines currently used in diagnostic laboratories. In our study, sample cross-contaminations, as previously discussed by Jacot et al. [10] with reference to low frequency mutations, can be reasonably ruled out as co-infections were suggested by the SNP profiles and confirmed by WGS using three sets of primers, each time in independent experiments using new RNA extracts. The relevance of low frequency clade-defining mutations was verified on the Integrative Genomics Viewer for all samples in this study. Moreover, in other samples not suspected of co-infection and analysed in the same WGS runs, the absence of low frequency 21J and 21K clade defining mutations was also checked.

The clinical presentation and vaccination status of co-infected patients were unremarkable. Analysis of follow-up samples showed that Delta or Omicron can outcompete the other, as previously reported although in immunocompromised patients [8]. Complementary analyses, such as neutralizing antibody levels against the two variants, could help to better understand if the ability of variants to evade immunity after vaccination could explain why one variant outcompetes the other after co-infection.

The evidence of Delta/Omicron co-infections makes it possible for recombination events to occur, a widespread evolutionary mechanism among RNA viruses including coronaviruses [11–13]. In our study, the distribution of VOC defining mutations for the P6 sample was not homogenous along the genome, suggesting a possible recombination event which need to be confirmed by virus isolation and long read sequencing. As of 23 February 2022, four French SARS-CoV-2 sequences were identified as one recombinant [6], which sequence is not genetically related to the one of the P6 sample (data not shown). Genetic recombination may generate new variants with unpredictable epidemic or pathogenic properties. This underlines the need for continuous efforts to maintain an effective genomic surveillance of SARS-CoV2 infections and a re-enforced surveillance of co-infection with emerging variants.

## Data Availability

All data produced in the present work are contained in the manuscript.

## Ethical statement

This study was based on national surveillance data. Informed consent was obtained from all patients. The study was approved by the review committee of the University Hospital of Clermont-Ferrand, France 2022/ CE06.

