## Supplementary figure 1 for "Evidence of co-infection during Delta and Omicron variants of concern co-circulation, weeks 49-2021 to 02-2022, France"

Figure S1

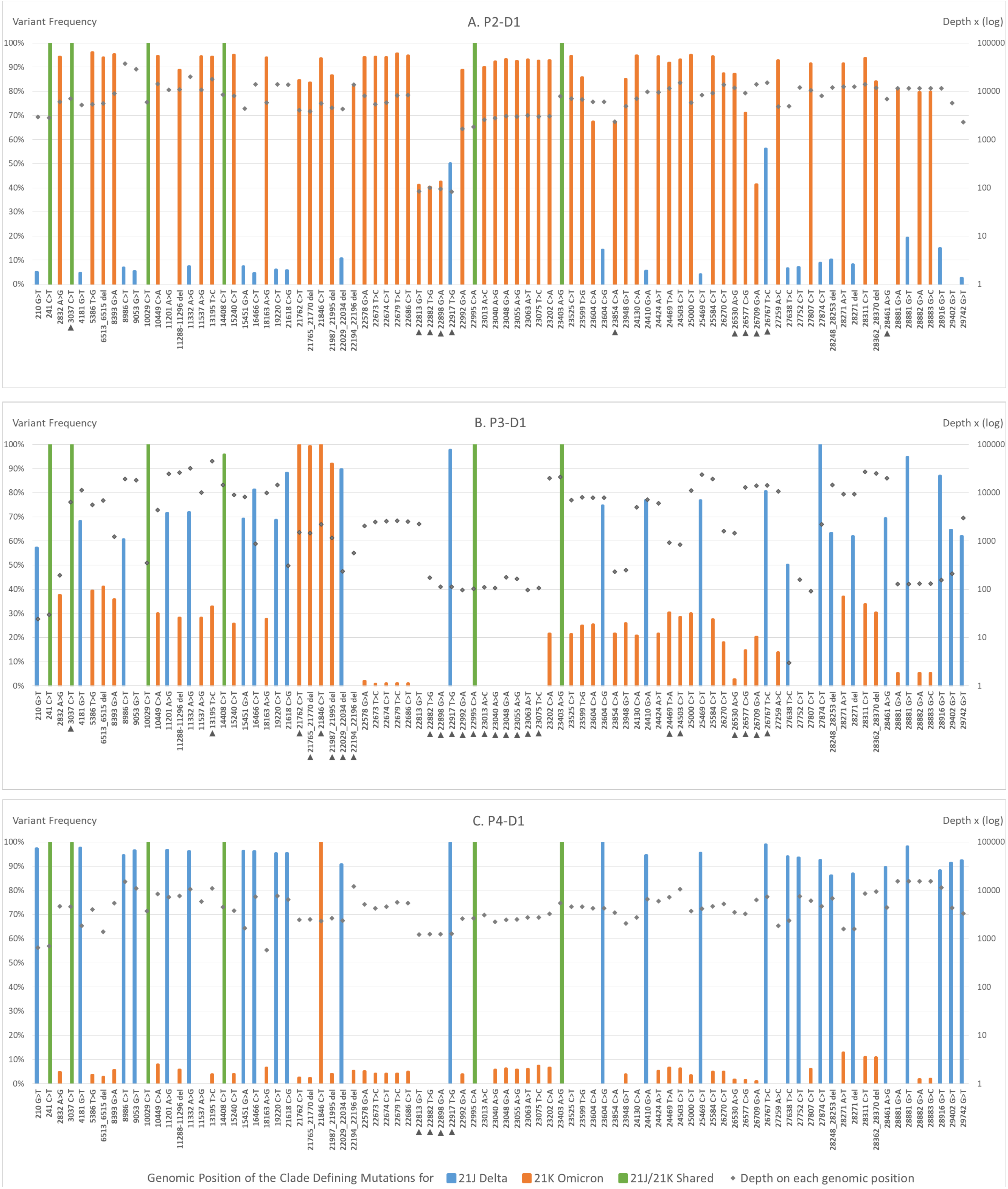


**Supplementary Figure S1**. Representation of Variant Allele Frequency clade defining mutations for 21J Delta and 21K Omicron along the SARS-CoV-2 genome for patients P2, P3 and P4.

All 21J and 21K clade defining mutations obtained by WGS (with Artic v4 primers for all patients or Artic v3 for sample P3) along the genome are presented for P2-D1 (A), P3-D1 (B) and P4-D1 (C). Mutations are represented with the VAF on the first Y axis and the depth of each genomic position on the secondary Y axis. If the mutation was not present in the vcf file, the value was set to zero. ►: clade defining mutations or deletions excluded for the proportion of co-infection estimation
