## Supplementary figure 2 for "Evidence of co-infection during Delta and Omicron variants of concern co-circulation, weeks 49-2021 to 02-2022, France"

Figure S2

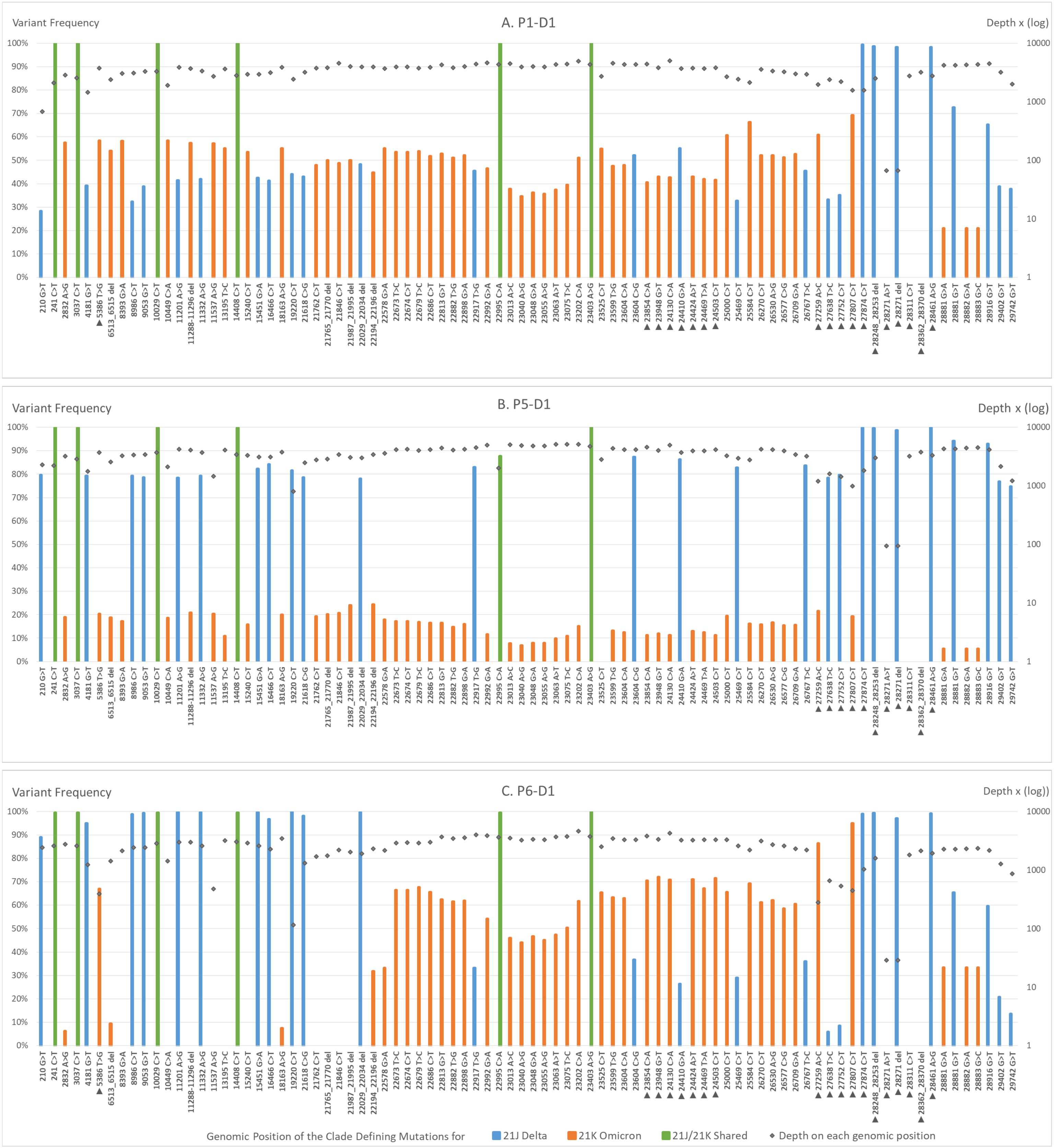


**Supplementary Figure S2**. Representation of Variant Allele Frequency clade defining mutations for 21J Delta and 21K Omicron along the SARS-CoV-2 genome using the Midnight 1200 primer set.

Co-infections for patients P1-D1 (A.) , P5-D1 (B.) and P6-D1 (C.) were confirmed with another WGS method using independent RNA extraction and xGen™ SARS-CoV-2-Midnight-1200 Amplicon (IDT) sequencing on a NovaSeq Illumina Platform (paired end reads of 100bp). Similar distribution of clade defining mutations to the ones obtained with the artic primer sets were observed except for the 3' end of the genome for which Omicron detection is known to be affected by mutations on primer sites with the Midnight v1 primer set [5]. Percent of co-infections, calculated with the third method described in the manuscript, provided similar results: P1-D1: 43.7% ± 10.6% Delta and 48.3% ± 10.4% Omicron, P5-D1: 82.0% ± 5.0% Delta and 14.5% ± 5.2% Omicron, P6-D1: 72.1% ± 33.0% Delta and 38.8% ± 26.8% Omicron. ►: mutations not included in the co-infection percent estimation as their amplification can be affected by primer mismatches due to other genomic viral mutations.
